# Assessment of N95 respirator for reuse after sterilization: filtration efficacy, breathing resistance, quality factor, chemical structure and surface charge density

**DOI:** 10.1101/2021.05.25.21257801

**Authors:** Amit Kumar, V. Subramanian, J. Shailesh, B. Venkatraman

## Abstract

In the present ongoing pandemic, the N95 respirator is an essential protective barrier to suppress the spread of the SARS-Cov-2 virus and protect the frontline worker from exposure. The N95 respirators are meant for single usage; however, they can be used after decontamination in-light of the economy and shortfall in availability. At this juncture, the respirators performance after various types of sterilization and usage condition is required to be analyzed in detail. With this motto, this work has proceeded. The filtration efficiency, pressure drop, and quality factor of the respirator are evaluated for two face velocities (5.8 and 26.4 cm/s) following different sterilization methods. Sterilization techniques used here are dry air oven heating, gamma irradiation, and immersing in a 10% concentration of liquid hydrogen peroxide. The particle filtration performance and electrostatic surface charge density measurement are used to determine the facemasks efficacy after sterilization. The methods recommended to sterilize N95 masks without affecting their performance are (i) using dry air heat at 80°C and (ii) H_2_O_2_ soaking. The highest reduction in filtration efficiency is observed to be 30-35% after gamma irradiation due to a change in the electrostatic properties of the respirator layers. However, the filtration efficiency does not change significantly for other sterilization methods despite a change in charge density, but there is no direct correlation with filtration efficiency. Electrostatic charge measurement of the filtration layer is a crucial indicator of filtration efficiency degradation. Policymakers can use these data during potential future N95 shortage to assess the viability of sterilization methods.

## 1.0 Introduction

Personal Protective Equipment (PPE), such as respiratory facemasks, offer protection against the spread of COVID-19through droplet and aerosols transmission (Wang and Du, 2020). When an infected person speaks, sneezes and coughs, the virus-containing droplets (> 5.0 µm) and aerosols (< 5.0 µm) will be excreted, remain suspended for up to a few hours and travel a longer distance on the air (Prather et al., 2020 and Guzman, 2020). N95 is essential PPE to protect the wearer with at least more than 95% filtration efficiency for Most Penetrating Particle Size (MPPS) 0.1 - 0.3 µm and higher efficacy for those particles smaller or bigger than MPPS (Qian et al., 1998 and Centers for Disease Control and Prevention, 2020). Hence, N95 offers excellent protection when they correctly seal over the face, where filter fabric made of Nylon, cotton, polyester, and polypropylene (Chawla, 2016). Even though the N95 facemask designed for single use, the recent pandemic crises made it necessary to seek emergency alternatives. Toward this crisis, facemasks are being reused multiple times after sterilization with the appropriate method without compromising effectiveness.

Several non-traditional sterilization/decontamination methods have evaluated with various successes (Hearn et al., 2020 and Kumar et al., 2020 c). Many potential sterilization methods have been explored already, and they are based on chemical and physical processes. The chemical methods include immersing the mask with liquid/ vapour hydrogen peroxide, chlorine dioxide, ethylene oxide, bleach, alcohol, soap solution and ozone decontamination. The techniques such as soaking/dipping in alcohol, chlorine, soap, and bleach damage the fibers of facemasks and significantly degrade respirators filtration ability (Peltier et al., 2020 and Lin et al., 2017 a). The liquid/ vapor hydrogen peroxide and ethylene oxide are reported to preserve filtration characteristics, but it needs complicated equipment, lengthy procedure, low throughput and trained human resource (Viscusi et al., 2009). The physical methods include dry/steam heat treatment, microwave oven, hot water, UV light sterilization, electron beam and ionizing radiation(Kumar et al., 2015, Liao et al., 2020 and Wang et al., 2020). Although UV radiation does not affect the filtering efficiency, they seldom penetrate through the respirator’s filter layers, resulting in partial sterilization (Fischer et al., 2020). Sterilization by ionizing radiation and electron beam well-known technology with large throughput for PPE and other medical types of equipment; however, recent studies show that the N95 facemasks filtration efficiency significantly deteriorates after gamma sterilization (Kumar et al., 2020 a and b, Pirker et al., 2021). The dry/ steam heat treatment has emerged as a promising and straightforward reprocessing method of respirators with good scalability and uniformity disinfection (Xiang et al., 2020; Liao et al., 2020). The steam sterilization by autoclave found helpful as far as filtration efficiency, but it has slightly melted the N95 mask plastic cord (Kumar et al., 2020 c). However, understanding the physical and chemical changes induced by the above various treatments remains unexplored or incomplete in a few cases.

Various decontamination processes on the physical and chemical change in the fiber of N95 respirator have been studied in the current work. The sterilization methods used in this work are (i) ionizing radiation (15 and 25 kGy), (ii) dry heat (30 and 60 minutes) and (iii) soaking in 10% concentration of hydrogen peroxide for 30 minutes. The respirators first tested for filtration performance, then were sterilized using the methods mentioned above and tested for filtration efficiency. Here, we carefully studied the filtration efficiency and facemasks charge density to detect a relation to the electrostatic filtration before and after various sterilization. In addition to that, the chemical change to the facemask’s fiber was examined after various decontamination. Most of the current works focus nearly exclusively on the respirator’s filtration performance (Jaung et al., 2020, Cai et al., 2020, Liao et al., 2020, Xiang et al., 2020, Anderegg et al., 2020 and Fischer et al., 2020). The current work aimed to investigate the effectiveness of the N95 respirators after disinfected by various methods and, most importantly, the origin cause for degrading respirator performance. The detailed experimental materials and methods, data analysis, results and discussion, conclusion are described in this article.

## 2.0 Materials and Methods

### 2.1 Protective respirator

The certified N95 respirators (Magnum) by the National Institute for Occupational Safety and Health (NIOSH) is used in the present studies. The N95 respirators (Make: Magnum Health & Safety Pvt. Ltd., India) is a pouch/ duckbill type, and an optical image is shown in Fig. 1. The Filtration Efficiency (FE) of NIOSH certified respirator is more than 95% for most penetrating particle size (Zhuang et al., 2005).

**Fig.1.**
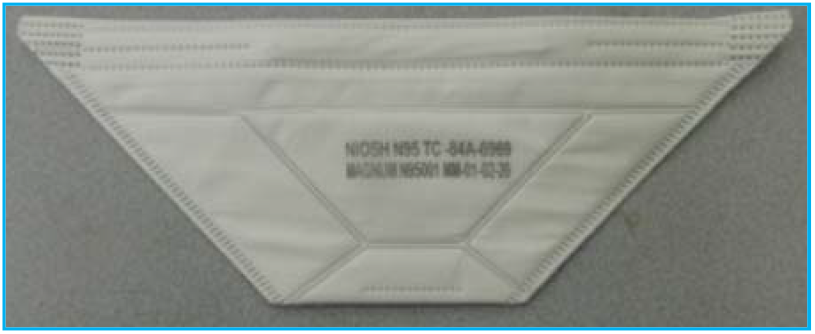
An optical image of certified N95 respirators used in the present study.

### 2.2 Material test setup

The experimental setup consists of an aerosol’s generator, aerosols diagnostic instruments and a differential pressure monitor. The respirators are evaluated in the HEPA Filter Testing Laboratory, Indira Gandhi Centre for Atomic Research (IGCAR). The facility consists of a stainless-steel cylindrical duct which has provision for fixing the facemask without any leakage. The experimental setup details, necessary data acquisition systems, and FE evaluation of masks for ambient and laboratory-generated aerosols explained in earlier work (Kumar et al., 2020 a & b).

### 2.3 Sterilization/decontamination methods

The following sterilization methods are employed viz. ionizing irradiation, dry heat, and soaking in H_2_O_2_ liquid in the present study. The respirators were irradiated with ionizing radiation of two doses, 15 and 25 kGy, using gamma chamber GC 500 having a dose rate of 1.80 Gy/h. The typical radiation dose for medical equipment sterilization (24kGy) and deactivation of virus (SARS-1) is around 20 kGy (Feldmann et al., 2019). The details of the gamma irradiation chamber are described in Kumar et al.,2020 b. Before irradiation, each mask was packed into an airtight plastic bag to prevent any contamination during irradiation. The dry heat decontamination of respirators was carried out with two different time durations of 30 and 60 minutes using a hot air oven (Labtherm Scientific Products, Sr. No.0013) with air temperature ranging from 80±3°C. The temperature and time of sterilization were chosen to deactivate SARS-COV-2 without damaging the facemask fiber integrity (Pascoe et al., 2020). The soaking treatment was carried out with a 10% concentration of H_2_O_2_ (diluted with distilled water) for 30 minutes. Special care has been taken in each decontamination process not to bend or damage the respirator to avoid the reduction in filtration efficiency, decreasing surface charge density and mechanical properties. An optical image of the gamma irradiation chamber, hot air oven, and H_2_O_2_ sterilization are given in supporting information (Fig.S1). The facemask, which is liquid submersion (soaking in H_2_O_2_) has been dried overnight before performing the FE testing.

### 2.4 Data acquirement and analysis

The *FE* of the respirator is calculated before and after decontamination using the following formula:

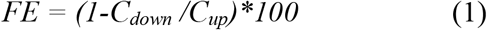

where *C*_*up*_ and *C*_*down*_ are the average aerosols concentration in counts/liter of upstream and downstream. The sampled data has been recorded for about two □ three minutes in 6 seconds frequency of each respirator from aerosols spectrometer (total sample record 20□30). *FE* estimated by taking the average of the collected data (upstream and downstream) for each mask tested period. *FE* averaged for all similar type masks and tested conditions (face velocity and sterilization methods). The uncertainty associated with *FE* of respirator has been calculated based on Type A and Type B uncertainty (Kumar et al., 2020 b). After that, the combined uncertainty has been assessed and then expanded uncertainty resulting from combined uncertainty by multiplying the coverage factor (95% confidence level).

### 2.5 Surface charge measurement

The electrostatic surface charge density is determined by measuring the surface charge of the respirator. The Keithley electrometer (Make: M/s. Tektronix USA and Model: 6517B) used for charge measurement of the respirators, and the schematic presentation is given in Fig.2.The electrometers input is a three-lug (Triax) connector, with the red-colored innermost wire (input high) is the charge-sensing terminal. As shown in Fig. 2, the charge-sensing terminal is connected to the metal surface (Copperplate). The metal noise shield (another copper enclosure) connected with black wire (input low), and the third wire (green color) is grounded. Both copper plate and noise shield are electrically isolated by introducing an insulating Teflon surface in between. The charge measuring range by Keithley electrometer is 0.002 to 2 µC with an accuracy of 0.4% of the measured value. The test sample is a piece that has an area of 10 cm^2^ (4 cm x 2.5 cm) cut from separated each mask layer. The surface charge on each layer for the respirator has been measured and recorded.

**Fig.2.**
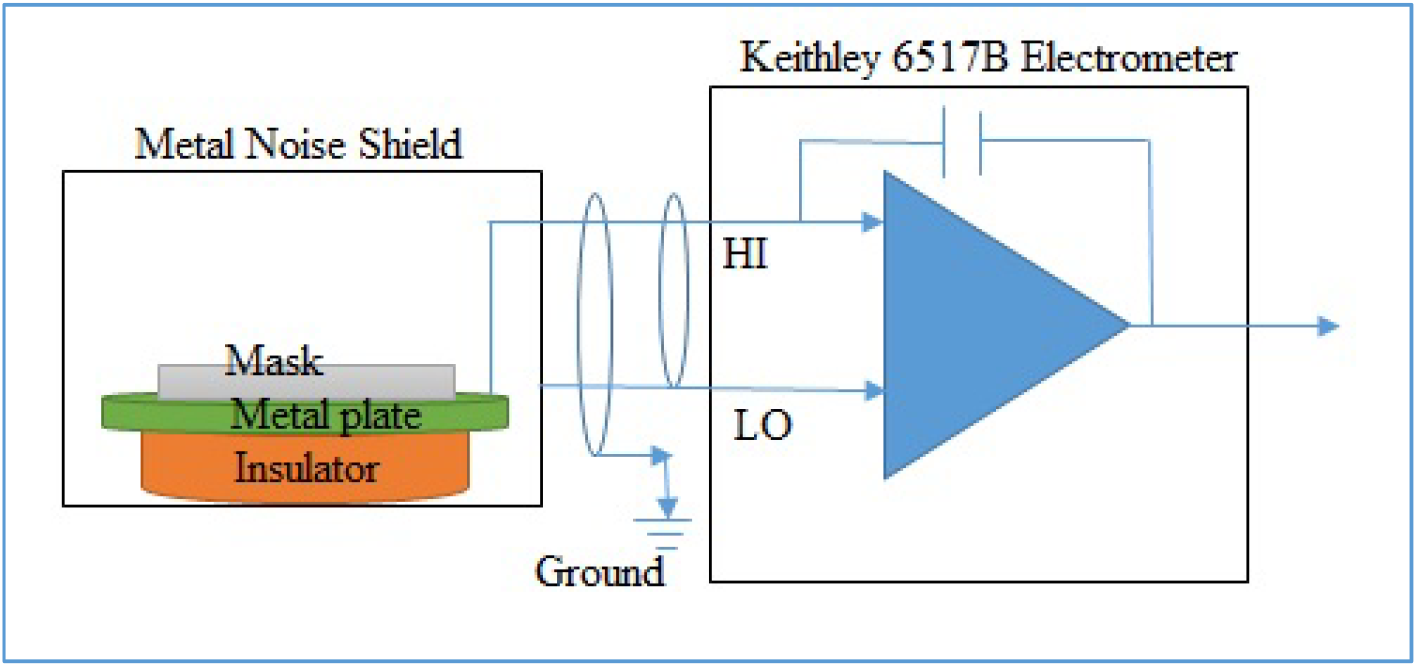
The schematic presentation of the surface charge measurement setup.

## 3.0 Results and discussion

### 3.1 Differential pressure drop across the facemasks

The measured pressure drops (ΔP) across the respirators before and after sterilization is given in Table 1. As expected, the ΔP across respirators increases with an increase in face velocity. The average pressure drop of control respirators is 31.38±2.16 and 258.90±4.61 Pascal (Pa) for low face velocity and high face velocity correspondingly. The ΔP across respirator slightly decreased for low face velocity after dry heat and H_2_O_2_ sterilization. The ΔP almost remained the same after ionizing irradiation; it slightly increased after dry heat sterilization and dropped after H_2_O_2_ disinfection for high face velocity. The pressure resistance indicates the condition for usage during breathing (even for high face velocity) is in the accepted range, 345 and 245 Pa for inhalation and exhalation resistance limit, respectively (Lin *et al*., 2020).

**Table 1.** Filtration efficiency, pressure drop and quality factor of certified respirator for two face velocity, control, and after decontamination.

| Sterilization<br>methods | Face velocity: $5.8 \pm 0.2\text{ cm/sec}$ | | | Face velocity: $26.4 \pm 0.9\text{ cm/sec}$ | | |
| --- | --- | --- | --- | --- | --- | --- |
| | Mean | $\Delta P \pm \text{Error}$ | $Q_f \pm 0.001$ | Mean | $\Delta P \pm \text{Error}$ | $Q_f \pm 0.001$ |
| | FE ± SD (%) | (Pa) | ($\text{Pa}^{-1}$) | FE ± SD (%) | (Pa) | ($\text{Pa}^{-1}$) |
| **Control** | $99.73 \pm 0.08$ | $31.38 \pm 2.16$ | 0.188 | $95.31 \pm 0.50$ | $258.90 \pm 4.61$ | 0.012 |
| **Heat (30 min)** | $99.30 \pm 0.52$ | $30.60 \pm 2.26$ | 0.162 | $97.65 \pm 0.51$ | $264.98 \pm 3.73$ | 0.014 |
| **Heat (60 min)** | $99.34 \pm 0.48$ | $30.65 \pm 2.25$ | 0.164 | $97.55 \pm 0.46$ | $265.21 \pm 3.69$ | 0.014 |
| **$\text{H}_2\text{O}_2$** | $99.15 \pm 0.38$ | $30.40 \pm 2.45$ | 0.157 | $95.88 \pm 0.61$ | $238.11 \pm 3.53$ | 0.014 |
| **15 kGy** | $68.69 \pm 0.35$ | $30.89 \pm 2.64$ | 0.038 | $60.92 \pm 1.43$ | $257.71 \pm 3.59$ | 0.004 |
| **25 kGy** | $70.44 \pm 0.54$ | $30.90 \pm 2.65$ | 0.040 | $62.66 \pm 0.39$ | $257.75 \pm 3.63$ | 0.004 |

**Table 2.** Percentage change in FE and σ of respirator for two face velocity after various decontamination.

| **Sterilization methods** | **U (5.8 cm/sec)** | **U (26.4 cm/sec)** | **$\Delta \sigma$ (%)** |  |  |
| --- | --- | --- | --- | --- | --- |
|  | **$\Delta FE$ (%)** | **$\Delta FE$ (%)** | **Outer** | **Filter** | **Inner** |
| **Heat (30 min)** | 0.43 | -2.46 | -6.07 | 22.01 | 3.05 |
| **Heat (60 min)** | 0.39 | -2.35 | -20.55 | 10.83 | 7.12 |
| **H<sub>2</sub>O<sub>2</sub>** | 0.57 | -0.59 | -24.52 | -55.13 | -48.98 |
| **15 kGy** | 31.12 | 36.09 | 135.92 | 117.78 | 76.99 |
| **25 kGy** | 29.36 | 34.26 | 155.08 | 155.12 | 71.80 |

### 3.2 Evaluation of filtration efficiency of masks for aerosol optical size > 0.3µm

FE of control facemasks was determined for the particle of optical diameter >0.3 µm, and two face velocities (Table 1). As expected, the *FE* of certified N95 masks is greater than 95%, and FE is more for low face velocity and less for high face velocity. The mean *FE* of control masks is 99.73±0.08 for low face velocity and 95.31±0.50for for high face velocity. The *FE* of respirators remained the same after dry heat and H_2_O_2_ sterilization for low face velocity. The FE slightly increased after dry heat for high face velocity and remained the same after H_2_O_2_ sterilization. The *FE* significantly decreased after gamma decontamination for both flow conditions. The certified face masks (N95) have an electret fibers filter that is efficient at low airflow velocity rather than high airflow velocity (Colbeck and Lazaridis, 2014).

### 3.3 Quality Factor (*Q*_*f*_)

A commonly used relation for filtration quality factor to determine the performance of the respirator defined as (Huang et al., 2013, Lin et al., 2017 a, b):

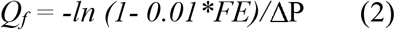

Where *FE* is in % and *ΔP* is in Pa. A higher *FE* and lower *ΔP* yield a higher filtration quality factor, i.e., better performance of the respirators. When the *FE* is ≥ 95%, the quality factor is more than0.031 Pa^-1^and 0.022 Pa^-1^for inhalations and exhalation resistance limit at 345 and 245 Pa pressure drop. The quality factor of the facemask for both face velocity is included in Table 1. The *Q*_*f*_ of control and decontaminated respirators ranges from 0.188 – 0.157 Pa^-1^for 5.8 cm/s face velocity and 0.012 – 0.014 Pa^-1^for 26.4 cm/s face velocity except gamma radiation (0.04 and 0.004 Pa^-1^for low and high face velocity).The *Q*_*f*_ values of respirators at low face velocity and after sterilization are found more than defined values for inhalation and exhalation resistance. However, it is found less than the defined limits for high face velocity due to high-pressure drop across the mask (except gamma sterilization). In the case of ionizing radiation decontamination, the *Q*_*f*_ value is less due to *FE* reduction.

### 3.4 Structure component of respirator

The Magnum N95 respirators were disassembled to understand their structural components. The masks made of three primary layers: outer, filter and inner layer. The filter layer of the respirator is sandwiched between the outer and inner layer. The respirators used spun-bond Polypropylene (PP) fabric for the outer and inner layer and melt-blown PP fabric for the filter layer. The filter layer of respirators composed of two layers. There is no observable visual change was found after various sterilization processes. However, after H_2_O_2_ sterilization, the printed matter on the surface of facemasks is appeared to be faded.

### 3.5 Chemical change of respirators fiber

The N95 respirator comes in many brands and models that use different materials and method to meet industrial filtration requirements. Most of the N95 respirators use a melt-blown electret material that employs the electrostatic forces to increase the filtration for MPPS. Ionizing radiation and other sterilization methods are known to modify the properties and structure of the polymer. They can adversely affect filter fiber in several ways, such as enhancing polymer oxidation, chain scission, cross-linking, and charge neutralization of electret material (Viscusi et al., 2009; Harrell et al., 2018). To check any significant change in the chemical structure of PP fibers has been caused after sterilization. The respirators have been analyzed before and after decontamination by Attenuated Total Reflectance-Fourier Transform Infrared spectroscopy (ATR-FTIR). All the spectra of respirator layers were recorded from 4000 to 500 cm^-1^ wavenumber using FTIR spectrometer (Make: ABB and Model: MB 3000) with a deuterated triglycine sulphate detector and analysed by Horizon MB Software. All measurements were made using 50 scans and 4 cm^-1^ resolution. Typical FTIR spectra of respirators with and without decontamination are shown in Fig. 3. The ATR-FTIR spectrum of the control respirator exhibits four IR peaks in the range of 2750 to 3000 cm^-1^ due to stretching vibrations of CH_2_ and CH_3_ groups. Peaks at 2950 and 2871 cm^-1^ are due to CH_3_ asymmetric and symmetric stretching vibrations, respectively, while peaks at 2908 and 2833 cm^-1^ are due to CH_2_ asymmetric and symmetric vibrations, respectively (Morent et al., 2008 and Ullah et al., 2020).

**Fig.3.**
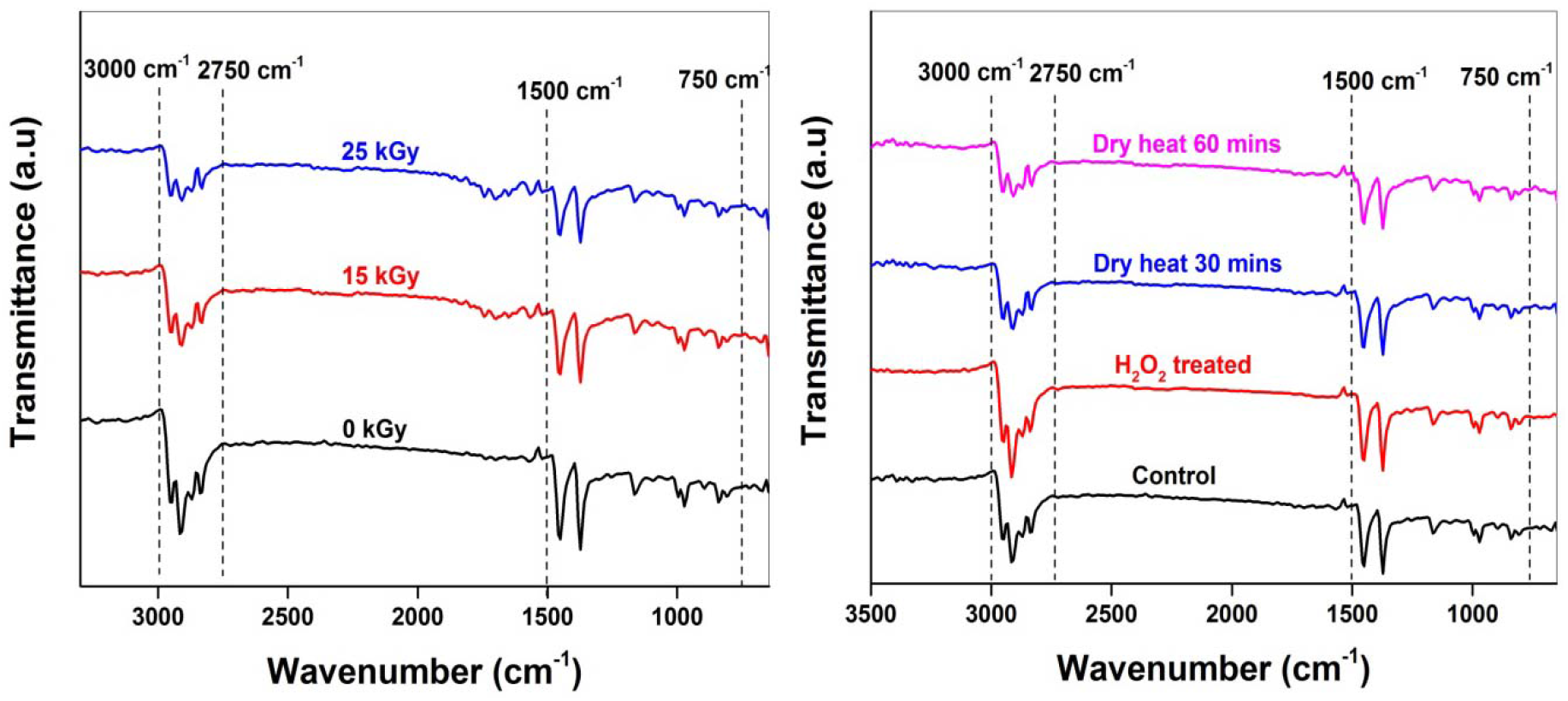
A typical ATR-FTIR spectra in the transmission mode of the filter layer of control and decontaminated respirators.

The ATR-FTIR spectrum of respirator also contains two intense peaks and various small peaks in 1500 to 750 cm^-1^. The peak at 1457 cm^-1^ is attributed to CH_3_ asymmetric deformation or CH_2_ scissor vibrations, while the peak at 1373 cm^-1^ is due to CH_3_ symmetric deformation vibrations (Morent et al., 2008). Absorption peaks displayed at 1162, 997, 971 and 899 cm^-1^ are assigned to CH_3_ rocking vibrations, while peaks at 843 and 805 cm^-1^ are assigned to CH_2_ rocking vibrations. Also, the peaks at 1162, 971 and 899 cm^-1^ can be given to C-H wagging, C-C asymmetric stretching and C-C symmetric stretching vibrations, respectively (Morent et al., 2008, Gopanna et al., 2019). It is clear from the ATR-FTIR spectrum of respirators that the filter layer is made of polypropylene fiber. The peak position and number of peaks remain the same before and after various sterilization methods (Fig 3). It can be concluded that gamma irradiation and other sterilization methods have not caused any significant changes in the molecular structure of PPs.

### 3.6 Electrostatic surface charge density (σ)

The electrostatic charge of respirators is measured before and after decontamination to evaluate the effect of surface charge density on *FE*. The average surface charge of a full respirator (without cutting and separating each layer) is measured and summarized in table S1. The average surface charge on the control respirator is + 0.841±0.441, which is in line with other literature (Hossain et al., 2020). The polarity of the surface charge on the respirator changed after sterilization; however, the average charge magnitude has increased despite decontamination methods. The variation in the average surface charge of respirators from a control mask ranges from 351-992%. The highest variation observed after gamma sterilization, while the lowest variation in dry heat for 60 minutes. Here, we observed that the full respirators charge measurement does not convey any correlation with filtration performance. However, *FE* has decreased after gamma sterilization, but the average charge on the whole surface increased.

To investigate more insightful information on the surface charge on each layer of respirator has been investigated. As explained earlier, the respirator structure consists of three layers. Surface charge measurement is carried out by separating each layer, and care has been taken not to damage the filter media, which would alter the electrostatic charge measurement. According to Lustig et al., 2020, only the filter layer acts as a filter for blocking particles with maximum filtration efficiency; other layers support the filter layer and filter large particles (Lustig et al., 2020). Hence, each layers electrostatic charge measurement is carried out by cutting a sample area 10 cm^2.^ The electrostatic surface charge measured of each layer of respirators is summarized in table S2. At the same time, σ (nC/m^2^) is calculated from each respirator layers from measured charge and surface area and shown in Fig. 4.

**Fig.4.**
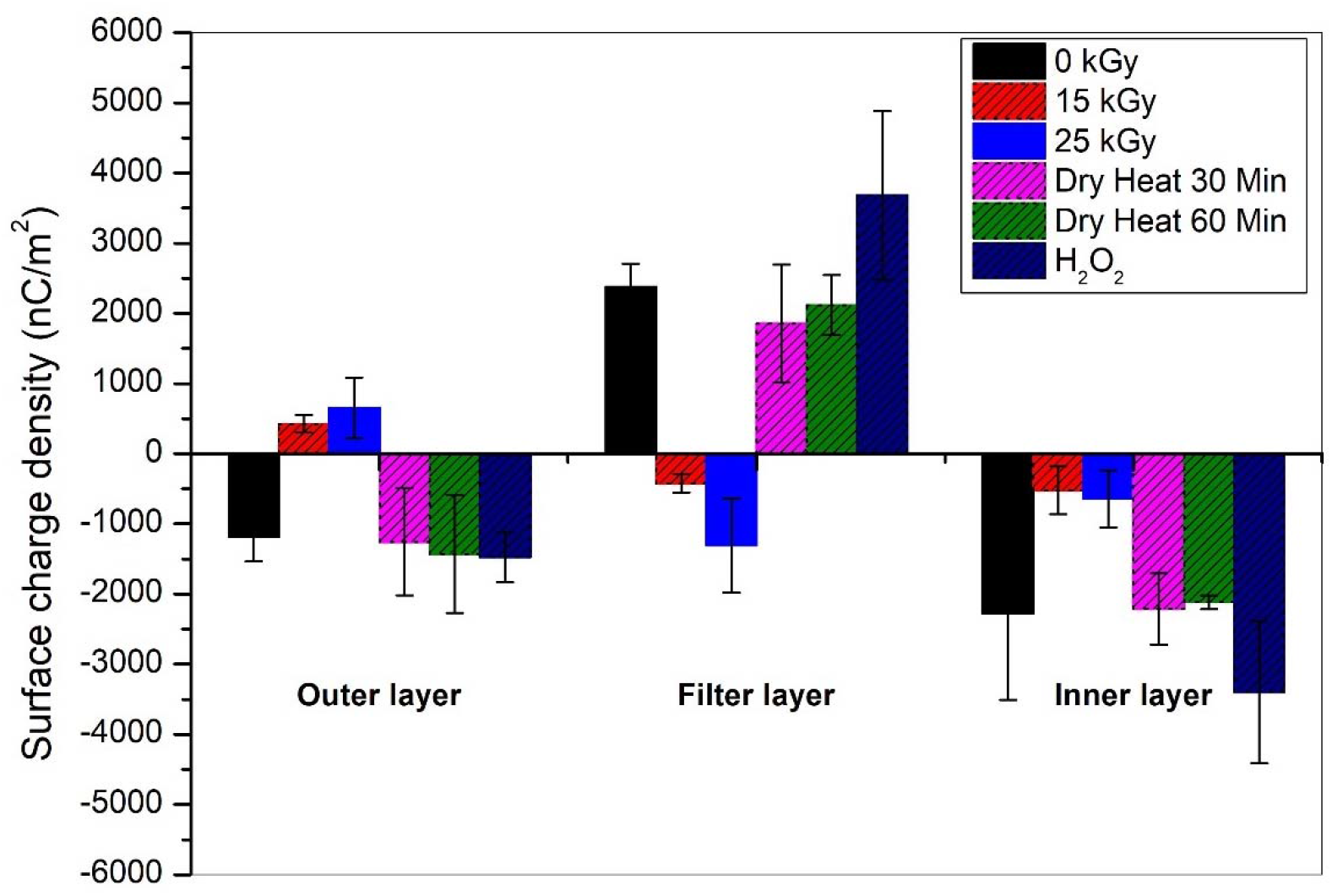
Electrostatic surface charge density on each layer of control and sterilized respirator.

The measured σ of control respirator is negative polarity for the outer (−1180±348 nC/m^2^) and the inner surface (−2279±1227 nC/m^2^), while for filter surface it is positive (+2374±335 nC/m^2^). The charge density imposed on all layers of the respirator has almost the same order. After sterilizing the respirator by dry heat and H_2_O_2_, the polarity of charge density remains similar, but the magnitude persists for dry heat and increases for H_2_O_2_. After gamma irradiation, the charge density on the respirator changed the polarity of the outer and filter layer but remains identical for the inner layer; however, after gamma irradiation, a reduction in charge density is found significantly large. The calculated σ is one order less than the reported values of N95 and K95 (Yim et al., 2020). The measured surface charge density has a significant statistical variation. The absolute values are uncertain; however, the substantial change in charge density detected qualitatively. The electrostatic charge of control masks showed that the filtration layer has a significant amount of surface charge. It appears that the filtration layer reduced its surface charge after sterilization (polarity has changed for gamma irradiation). There is a qualitative correlation between surface charge density and FE; masks with higher values of σ having higher *FE*. The present observation on electret electrostatic charge in line with other literature (Hossain et al., 2020, Yim et al., 2020and Pirker et al., 2020) shows that the variation is not consistent. Nevertheless, the finding is consistent with other studies that have observed the degradation in the FE of respirator associated with filter media electrostatic degradation.

Further, the filter layer composed of two layers, both layers carefully separated, and surface charge measured on both filter layers of respirators and given in Table S3. One filter layer is negative polarity, and the other is positive for the control respirator. After sterilization of the respirator, the polarity of negative charge layers has not changed, but the charge magnitude has been altered. The positive charge layer polarity also does not change except gamma irradiation (positive polarity change become negative polarity). The average charge variation on the filter layer of respirators from the combined filter layer (both filter layers are not separated) ranges from 15-56% except 25 kGy sterilization (−31%). The negative variation in average charge means the separated filter layer has given more measured surface charge, while positive variation means the measured surface charge less on separated filter layers.

### 3.7 Assessment of facemask effectiveness after sterilization

The percentage change in *FE* and *σ* after different sterilization from control facemasks summarizes in table 3. The *ΔFE* of the mask from control to dry heat and H_2_O_2_sterilization is less than 1% and -3 %for low and high face velocity. However, *FE* changes around 29 – 36% for gamma sterilization for both flow conditions. The *ΔFE* is more for gamma sterilization and further increases for higher face velocity. The change in surface charge density on the outer filter and an inner layer of masks from control to dry heat sterilization ranges from 3.05 to -20.55%, H_2_O_2_sterilization ranges from -24.52 to -55.13%, and gamma sterilization ranges from 71.80 to 155.12%. The most significant reduction in charge density observed after gamma sterilization is in correlation to reducing mask filtration efficiency. The *FE* does not affect much after dry heat and H_2_O_2_ sterilization despite the change in surface charge density.

It is known that aerosol filtration occurs by five basic mechanisms viz. gravitational settling, inertial impaction, interception, diffusion and electrostatic force (Hinds, 1999 and Vincent, 2007). The N95 mask consists of electrets fibers media capable of capturing and retaining fine particles through electrostatic and mechanical filtration processes (Myers and Arnold, 2003). The least efficiency is associated with particles in the range from 100□500 nm, more prominent for diffusion and lesser for interception; hence, the 95% efficiency achieved for this range of particles by electrostatic force and dielectrophoretic force. When the electret media modified its charges (surface charge density), the efficiency may also get altered. It is found from the present studies that each layer of respirator and filter layer itself has not only different charge density but also the different polarity. The filter layer is composed of opposite charge layers, which gives rise to a strong electric field gradient inside the filter layers. In the absence of a strong inhomogeneous electric field, the respirators filtration properties are reduced, which is observed in the ionizing radiation sterilization method.

## 4.0 Summary and conclusion

The certified N95respirator evaluated for particulate FE, pressure drop, quality factor, and various disinfection methods for two face velocities 5.8 and 26.4 cm/s. As expected, the pressure resistance increases with increasing face velocity; this reflects that it is harder to breathe through a facemask when the respiratory flow is large. The measured pressure drops during masks testing found to be the accepted range for the inhalation and exhalation resistance. The quality factor of facemasks is in the acceptable content except after gamma irradiation. The *FE* found to be more than 95% for the control respirator and both face velocity. The *FE* is similar before and after dry heat sterilization for 30 and 60-minutes duration and reduces to about 70% after gamma sterilization for low face velocity and still lesser with higher face velocity. After H_2_O_2_ sterilization, the *FE* of facemasks remains unaffected. Thus, it is recommended that the N95 respirator can be sterilized a few times with dry heat and H_2_O_2_.

N95 masks, which made of electret filtering media, are not recommended for sterilization or decontamination by ionizing radiation; it will compromise the filtering efficiency. The FTIR spectra of facemasks after gamma irradiation and other sterilization have not caused any significant chemical changes, i.e., the molecular bond nature of the studied filter layer of respirators. The most significant reduction in charge density observed after gamma sterilization is in correlation with reducing respirator filtration efficiency. *FE* does not affect much after dry heat and H_2_O_2_ sterilization despite the change in surface charge density.

This study comprises electrostatic charge measurement to detect a change in FE after various sterilization methods. Some other test like quantitative fit test is intended for control materials, and it is limited in how much information they can give on the performance degradation of a respirator. They can only determine respirators fitness appropriate to the user; however, they cannot decide to what degree and why the mask is degraded. Most recent works focused on measuring the respirators surface potential and correlated to the filtration performance (Pirker et al., 2020 and Yim et al., 2020). Electrostatic charge measurement of the filtration layer is a crucial indicator of filtration efficiency degradation. The present study used to understand the degree of degradation and correlate with electrostatic charge properties on each respirator layers. However, it is worth noting that the lesser sample size and large variability in charge measurement made it challenging to obtain high certainty quantifiable measurements. Nevertheless, the finding is consistent with other studies that have observed the degradation in FE of masks associated with filter media electrostatic degradation. Further, the N95 respirators come in many brands and models that use different materials and fabrication methods to meet industrial filtration requirements. The electret filter charge measurement can be assessed for other brands of respirators and sterilization methods for their reusability. However, the behavior of charge density variation in the filter layer would be similar.

## Data Availability

The relevant data included in the manuscript and supporting information.

## 5.0 Acknowledgement

The authors acknowledge Shri S. Athamalingam, Associate Director, Health, Safety and Environmental Group, and Dr. R. Venkatesan, Head, Radiological and Environmental Safety Division for their encouragement and backing to carry out this work.

