## Supplemental Information for "Assessment of N95 respirator for reuse after sterilization: filtration efficacy, breathing resistance, quality factor, chemical structure and surface charge density"

### Materials and Methods

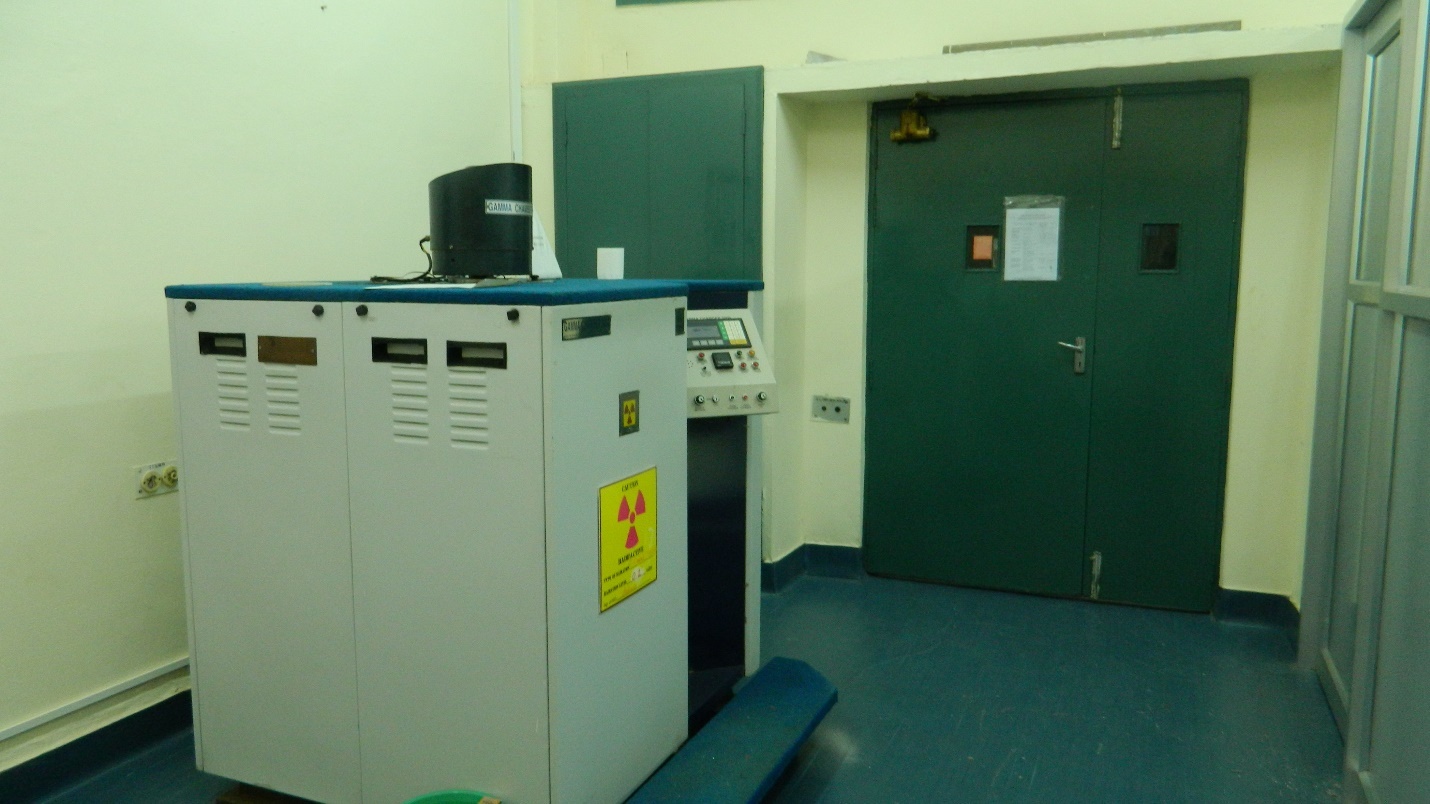

Fig. S1a. An optical image of gamma irradiator (Model and Make: GC500 and M/s BRIT, India) where the masks are irradiated using Co-60 source (gamma energy 1.17 and 1.33 MeV) for the desired dose level (15 and 25 kGy). The gamma irradiation chamber is cylindrical with approximately 17.2 cm in diameter, and 20.5 cm height and the dose throughout the chamber is uniform by using pencil sources (Kumar et al., 2020).

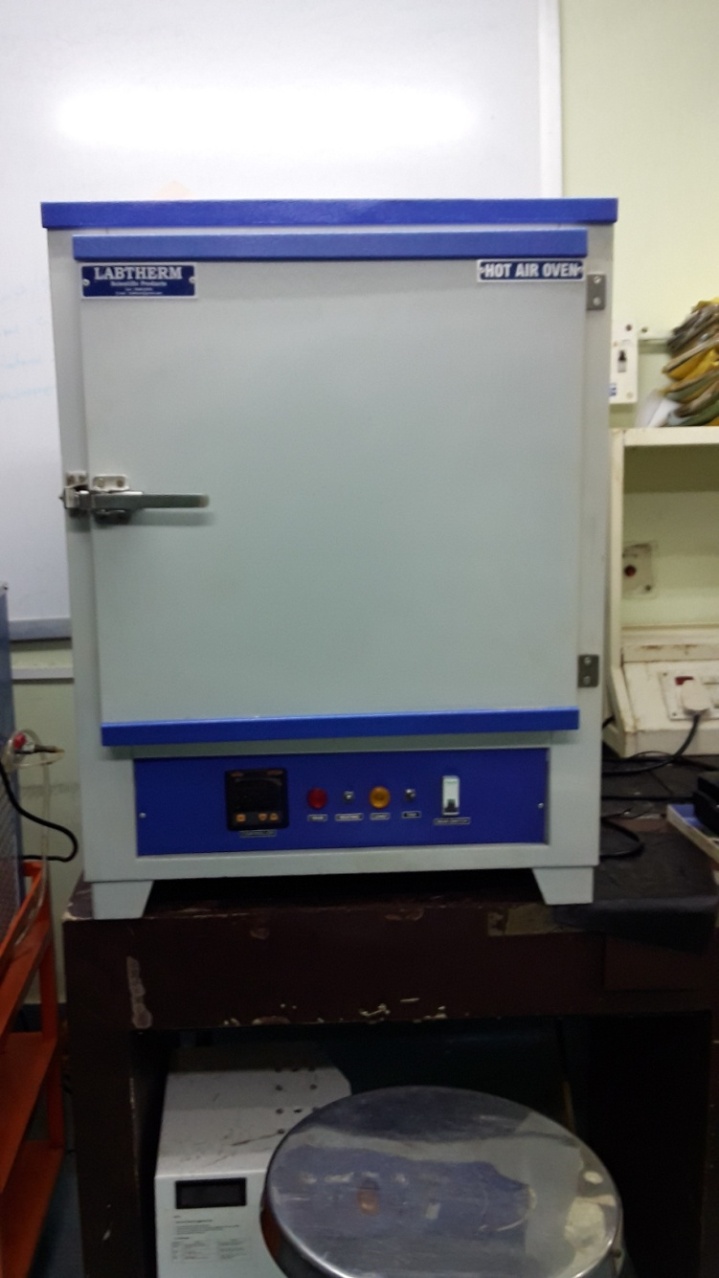

Fig. S1b. An optical image of hot air oven (Model and Make: and M/s Labtherm, India) where the masks decontaminated at 80±3^o^C air temperature for a period of 30 and 60 minutes.

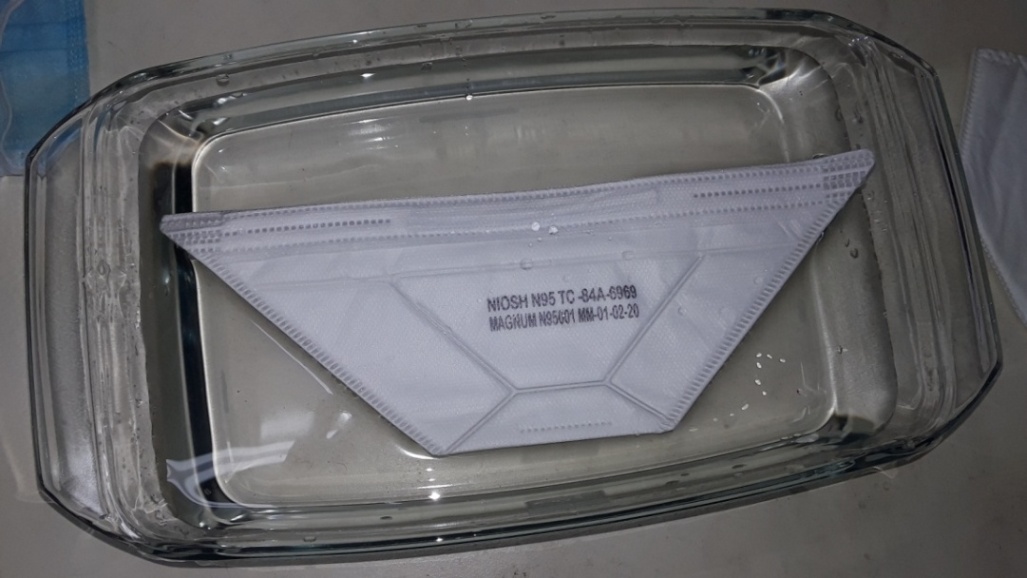

Fig. S1c. An optical image of magnum (NIOSH) respirator with H_2_O_2_ soaking for 30 minutes.

### Table S1. The measured surface charge of full respirators without cutting. It can be seen from the table that the polarity of the charge of respirator changed. The magnitude of the average charge on respirators has increased despite decontamination methods. The variation in average charge of respirators from control ranges from 351- 992%. The highest variation observed after gamma sterilisation while the lowest variation in dry heat sterilisation for 60 minutes.

| **Sterilisation methods** | **Average surface charge ± SD (nC)** | **Variation from control (%)** |
| --- | --- | --- |
| **Control** | 0.841 ±0.441 | NA |
| **Heat (30 min)** | -5.596±0.572 | 765 |
| **Heat (60 min)** | -2.113 ±0.805 | 351 |
| **H_2_O_2_** | -5.475 ±0.355 | 751 |
| **15 kGy** | -7.506 ±2.812 | 992 |
| **25 kGy** | -6.865 ±1.710 | 916 |

### Table S2. The measured surface charge on various layers of respirators. The measured charge of control respirator is negative polarity for the outer and an inner layer while for filter layer it is positive. After sterilisation of the masks by dry heat and H_2_O_2_, the polarity of charge remains the same but magnitude persist for dry heat and increases for H_2_O_2_. After gamma irradiation, the charge on a respirator changed the polarity of the outer and filter layer but remained the same for the inner layer; however, after gamma irradiation reduction in charge significantly large.

| **Sterilisation methods** | **Measured surface charge (nC) ± SD** | | |
| --- | --- | --- | --- |
|  | **Outer layer** | **Filter layer** | **Inner layer** |
| **Control** | -1.185 ±0.349 | 2.375 ±0.335 | -2.280 ±1.227 |
| **Heat (30 min)** | -1.257±0.766 | 1.852 ±0.841 | -2.210 ±0.508 |
| **Heat (60 min)** | -1.428 ±0.842 | 2.118 ±0.429 | -2.117 ±0.096 |
| **H_2_O_2_** | -1.475 ±0.355 | 3.684±1.202 | -3.396 ±1.104 |
| **15 kGy** | 0.426 ±0.125 | -0.422 ±0.133 | -0.525 ±0.340 |
| **25 kGy** | 0.653 ±0.434 | -1.309 ±0.669 | -0.643 ±0.406 |

### Table S3. The measured surface charge on both filter layers of respirators. The one filter layer is negative polarity, and the other is positive for control respirator. After sterilisation of respirator, the polarity of the negative charge layer has not to change. In contrast, charge magnitude has been changed except gamma irradiation (polarity also change of the second filter layer). The variation on the average charge on the filter layer of respirators from the combined filter layer (both filter layers are not separated) ranges from 15- 56% except 25 kGy sterilisation (-31%). The negative variation in average charge means the separate filter layer has given more measured charge while positive variation means the less measured charge on separated filter layers.

| **Sterilisation methods** | **Mean charge (nC)** | | | **Variation from control FL (%)** |
| --- | --- | --- | --- | --- |
|  | **FL-1** | **FL-2** | **FL-1+FL-2** |  |
| **Control** | -0.777 | 2.242 | 1.465 | 38 |
| **Heat (30 min)** | -1.837 | 2.650 | 0.813 | 56 |
| **Heat (60 min)** | -0.026 | 1.731 | 1.705 | 19 |
| **H_2_O_2_** | -0.215 | 2.045 | 1.830 | 50 |
| **15 kGy** | -0.286 | -0.072 | -0.358 | 15 |
| **25 kGy** | -1.055 | -0.670 | -1.725 | -31 |
